# Trends and Factors associated with use of Modern Contraception Methods in Rwanda: A comparative study using Rwanda Demographic and Health Surveys 2010, 2014/2015, and 2019/2020

**DOI:** 10.1101/2025.05.18.25327848

**Authors:** Boniface Nsengiyumva, Marcel Baziruwiha, Jean Marie Vianney Sindambiwe, Nzeyima Zephanie, Julienne Nyirarukundo, Joella Mukashyaka, Isabelle de Valois Ndishimye, James Humuza, Cyprien Munyanshongore

## Abstract

**Background:** Modern contraception methods remain the most reliable means to control and space births. However, their use remains low for various social and economic reasons, particularly in low- and middle-income countries (LMICs), including Rwanda. This study examined trends and factors associated with the use of modern contraceptive methods among women aged 15 to 49 years in Rwanda from 2010 to 2020, using data from the Demographic and Health Survey (DHS) data.

**Methods:** This study was a comparative study using cross-sectional surveys weighted data of 3 editions of the Rwanda Demographic and Health Surveys (RDHS); 2010, 2014/2015, and 2019/2020. The study population was composed by women aged 15 to 49 years enrolled in each DHS. The participants were selected using stratified, two-stage sampling. The study used descriptive statistics, bivariate analysis, and logistic regression to analyze the data.

**Results:** The study found that the proportion of women using modern contraceptive methods increased significantly from 24.8% in 2010 to 27.1% in 2014/2015 and 33.9% in 2019/2020 (p<0.001). In the three editions of Rwanda DHs, variations in factors influencing the use of modern contraception methods were observed. In 2010, higher likelihoods of using modern contraception methods were among women reporting 1 to 3 as ideal number children a couple should have (aOR: 3.6 with 95%CI: 1.2-10.5), in Muslim religion (aOR: 8.2 with 95% CI: 1.0-64.4), married or cohabiting relationship (aOR: 2.2 with 95%CI: 1.6-3.2), and among those living in Northern province (aOR: 2.1 with 95% CI: 1.2-3.5). However, women living in households with 11 or more members, those of the Protestant faith, and those whose husbands make decisions regarding contraception use were less likely to use modern contraceptive methods. Secondly, in 2014/2015, women who received FP information from a health facility were 1.6 (aOR: 1.6 with 95% CI: 1.2-2.1) times more likely to use contraception than those receiving the information from radio. Similar to the 2010 DHS, lower likelihoods were noticed among those living in households with 11 or more members. Lastly, in 2020, higher likelihoods of using modern contraception were among women aged 20 to 44 years (aOR ranging from 2.1 to 2.8) as compared to those aged 15 to 19 and 45 to 49 years old. Moreover, women who decide mainly on their use of contraception were more likely to use it compared to those whose husbands do or jointly decide. Like in 2010, likelihoods were also higher among women reporting 1 to 3 as an ideal of children than one should give birth to, and among married or cohabiting partners (aOR: 1.4 with 95% CI: 1.0-2.0). However, it was unclear why lower likelihoods were among women with increasing education level.

**Conclusion:** This study found that modern contraception use increased from 24.8% in 2010 to 33.9% in 2019/2020. However, disparities persist, particularly among women from larger households, Protestant backgrounds, and higher education levels. These findings highlight the need for targeted interventions to address barriers to contraceptive use, especially among adolescent girls, young women, and religious groups with lower usage rates.

## 1. Background

Universal access to reproductive health, as defined in the 2030 Agenda for Sustainable Development, requires increased uptake of modern contraception(1). However, despite substantial global progress in meeting family planning needs, significant global inequities in access to modern contraceptive methods remain remarkable (1). Globally, an estimated 874 million of the 1.9 billion women of reproductive age (15–49 years) use a modern form of contraception, while 92 million use a traditional technique. Since 1990, the number of people using modern contraceptives has almost doubled from 467 million to the present day. However, 164 million women still wish to postpone or prevent getting pregnant but do not use any form of contraception(2).

Although the modern contraceptive use has increased globally, the needs for family planning has been significantly expressed. The 2019 unmet needs were estimated at 10% while the modern contraceptive use stood at 76%. The progress report on 2030 agenda for Sustainable Development Goals (SDGs) informed that 190 million women of reproductive age expressing their need to stop or delay pregnancy were not using any Family Planning (FP) methods in 2019 (3). An enormous difference to satisfy unmet needs was observed between regions/countries and only 55% of needs met was observed in 42 countries(3).

Over all users, 45% adhered to permanent and long-term methods while 46% use short acting methods and 8% were users of traditional methods. Among all methods of contraception, female sterilization and male condom were the most used methods at 24% and 21% respectively(4). The United Nations Population Fund (UNFPA) world population dashboard reported that the contraceptive prevalence rate stood at 49% among all women and at 63% in married /union women. The modern contraceptive rate was 45% in all childbearing women and 57% in married/union women. The unmet needs were 9% in all women and 11 % in married/union women. The 2019 global political environment guaranteed access to sexual and reproductive health care (including FP), information and education at 73% (5).

UNFPA reported that in 2019, 59% of all women of reproductive age in more developed countries were using family planning methods and 71% of married/in union women used contraception. In this part of world, the modern contraceptive prevalence rate was at 53% in all women and at 62% in women living with their partners. Eighty-four per cent of developed countries had set rules and regulations that guarantee access to SRH care, information and education(5).

Even though people of developed countries have a high access to modern family planning methods, in some countries low adherence to FP methods had been reported and was linked to specific realities like promotion of legalized abortion, young population, expensive FP methods and mainly lack of information or knowledge on long lasting methods. Others reported side effects and fear of some methods as barriers to FP use (6–8).

In Low and Middle Income Countries (LMICs), policies favoring access to sexual and reproductive health (SRH) exist in more than 70% of least developed countries, however, the Contraceptive Prevalence Rate (CPR) is 32% in all women of reproductive age and 42% in married /union women of 15-49years old. Modern contraceptive methods were used by 28% of all women and 37% of married/ union women(5). Like in other parts of the world, individual/ demographic factors had been analyzed and are linked to the uptake of family planning methods. Women from this area were mostly subject to early marriage, which predisposed them to producing many children than expected. Unfortunately, young women (15-24 years) were identified as the least users of modern contraceptive methods. Tariku et al. revealed that only 14% of Ethiopian young women were using contraception. Additionally, social factors such as perceived social approval and perception of friend’s contraceptive practice were positively associated to their use(9).

Significant disparities are observed between Asian countries and within countries in regions(10). The contraceptive prevalence rate in this part of world is classified in 2 groups, where highest CPR is remarkable in China, Thailand, and Vietnam with 84.6%, 78.7 % and 75% respectively. The group with lowest CPR includes Pakistan Maldives and Samoa with 34.2%, 34.7%, 26.9% respectively (10). In China, intrauterine Device (IUD) and female sterilization are the most popular contraception methods, used at 40, 6% and 28,7% of all methods. The traditional methods are most common in Malasia17.9% Cambodia17.5% and Philippines (13.%)(10). Even though considered as threat to people’s right, strict policies promoted long term and permanent methods and limited access to other choices of FP. These policies significantly contributed to the increase of CPR in China(10).

Sub-Saharan Africa (SSA) is one of the regions that has not experienced a significant increase in the use of modern contraceptive methods in the past decade (27.8%)(11). In 2019, the United Nations reported that in 23 SSA countries, the satisfaction of FP demand was less than 50%. Countries like Chad, Somalia and South Soudan were still at less than 25% in satisfying the demand of modern contraceptive methods (3). Although there has been improvement in most countries towards reducing unmet needs, 15 countries from this region remain with more than 20% of modern FP unmet needs. Angola and Liberia were of the highest level of unmet needs with 26% and 25% respectively(3). Like in all other regions, young women remain the group with low modern contraceptive users, followed by women of 40 years and above(12).

According to the 2010 DHS report, the modern contraceptive prevalence was 25% in all women, and 45% among married women. The RDHS 2014/2015 claimed that the modern contraceptive use was 27.8% in all women of reproductive age, and 47.5% in married women. Comparing to the 2019/2020 DHS report, the prevalence of modern contractive use was reported to be at 35% in all women and 58% in married women(13), indicating a substantial increase in modern contraceptive uptake for the past 10 years among married women, while it is not the case in all women. The 2010 RDHS indicated that In all women, injectables were the most commonly used (14.6 percent), oral contraceptives (3.9 percent), implants (3.6 percent) and male condoms(1.8 percent)(14). For the 2014/2015 RDH, Injectable remained the most used FP method (in 24%), followed by pills and implants, each used in 8%. The 2019/2020 RDHS, indicated that implants became the most popular modern family planning method in all women, followed by injectable (9%) and pills (3.8%).

The contraceptive prevalence rate of the RDHS 2019/2020 was relatively highest in women of 30- 35 years (65.2%) and low (46%) in the group of 45-49 years. Fifty- three percent of young women (15-19 years) were using Family planning methods(12). Surprisingly this DHS revealed that 65% of rural women were using contraception compared to 61% in urban settings.

Rwanda has set different mechanisms to guarantee universal access to FP and ASRH services especially in modern family planning methods. These strategies include provision of FP methods by Community Health Workers, promotion and provision of IUDs immediately after childbearing, availing modern FP methods in all government health facilities, upgrading the reproductive health services package that respond to youth needs, creating secondary health posts that respond to FP needs in people served by ecclesiastic health facilities, building capacity of health care providers at all levels in FP, periodic review and updating the FP strategic plan, and many others. All of these resulted in tangible progress in family planning use(15). However, there is a need to quantify the effect through the analysis of the trend in the prevalence and a comparative analysis of factors backing or discouraging these interventions. In the same streamline, several analysis and studies were conducted in the framework of identifying the level of family planning use and its underlying factors. Most of these studies analyzed each single DHS. Beyond that, little has been done to compare the prevalence of modern family planning use and its underlying factors among childbearing women across different periods. Therefore, this study focused on the understanding of the tends and factors most influencing or impeding progress of modern contraceptive method use among Rwandan women. More specifically, this identified the trends and compare the prevalence of modern contraceptive methods use among Rwandan women aged 15-49 years for a ten-year period. It also determined and compared the factors associated with modern family planning use among Rwandan women aged 15-49 years, using the RDHS 2010, 2014/2015, and 2019/2020.

## 2. Methods and Materials

### Study design

A cross-sectional retrospective study using quantitative data collected from RDHS 2010, 2014/2015 and 2019/2020 was used to examine the trends in prevalence and factors associated with use of modern contraceptive methods among women of reproductive age in Rwanda.

### Study population

The target population for this study comprises of all women of reproductive age who used any contraceptive method between 2010and 2020. The sample frame was composed of 1,757,426 And 2,424,898 households from enumeration areas (EAs) defined in 2002 and 2012 respectively during the Population and Housing Censuses.

### Sample size

This study used exhaustive sample size from RDHS 2010, 2014/2015 and 2019/2020. Therefore, the sample size for this study included 13671 women from RDHS 2010, 13497 women from RDHS2014/2015 and 14634 women from RDHS 2019/2020 (16,17). This sample size was obtained using Taylor linearization method and jackknife duplication for variance parameter.

### Study Variables

The outcome variable for this study was a binary variable indicating whether a given woman used a modern contraceptive methods or not. Additionally, this study used 22 independent variables. These are age, marital status of woman, woman’s education, woman occupation, husband desire for more children, main source of FP information, decision maker for contraception use, knowledge of ovulatory cycle, age of woman at first sex, ideal number of children, children ever born, number of living children, partner’s age, partners, education, partner’s occupation, number of household members, religion, sex of HH head, province, type of residence, household wealth index, and owning a radio and/or a television.

### Data collection procedures

Data for this study came from the Rwanda Demographic and Health Surveys (RDHS) in 2010, 2015/2014, and 2019/2020. A sample of households from around the country was chosen. All women aged 15 to 49 who were regular inhabitants of the selected households or who stayed in the households the night before the survey were eligible to take part in the survey. The RDHS samples were stratified and drawn from the preceding census frames in two steps to choose samples separately in each sampling stratum and collect the data from selected households within an EA (18). Stratification was accomplished by dividing each district into urban and rural areas, with each entity serving as a sampling stratum.

### Data analysis

The univariate analysis was done using frequencies and percentages to describe all the study variables. The trends in the prevalence of modern FP use was illustrated using line chart and tested using Chi-square distribution. The second step was the bivariate analysis and consisted of assessing the pairwise association between the use of modern FP method and each of the independent variable. The logistic regression model was constructed to indicate the individual likelihood of each independent variable to influence the modern FP use. Thus, the level of statistical significance between the outcome variable and each of the independent variable was determined using crude odds ratios (COR) and Adjusted Odds Ratio (AOR) at 95% confidence level. Before any analysis, we generated weighting variables and these allowed us to adjust our analyses to sample design, sampling weight, clusters, and strata.

### Study tools

Basically, 3 questionnaires (household, women, and men) were used during the RDHS. For this study, women data sets were utilized to provide required information on modern contraceptive methods and other variables to achieve the study objectives. A pretest was undertaken for each RDHS to ensure the validity of the data collection instruments.

## 3. RESULTS

### Socio-demographic characteristics of the study subjects

For RDHS 2010, most of the study participants were youth (15-24years), 41.9% (5433 women), with primary education, 68.5% (9086 women), with home occupations, 67.1% (8880 women), and were not yet married/in union, 38.5% (5122 women). Majority of the surveyed women were from western province, 24.3% (3228women), and living in rural area, 85% (11271 women). Women’s source of information on FP was mainly radio, 44% (4521 women) and most of them do not know their ovulatory cycle, 87.6% (11620 women). The partner age was mostly ranging between 30 to 39 years, 35 % (2366 partners), with primary education, 66% (5383 partners), and majority of them 70.6% (5736 partners) had home occupation. The husband desire for children was mostly the same as one for the study participants; 58.8% (3935 husbands) and the decision for contraception use was in most of cases taken by both women and husbands, 88% (3092 couples) (Table 1).

**Table 1:** Characteristics for the study participants.

| Variable | RDHS 2010 |  | RDHS 2014/2015 |  | RDHS 2019/2020 |  |
| --- | --- | --- | --- | --- | --- | --- |
|  | Frequency | % | Frequency | % | Frequency | % |
| **Age (in years)** |  |  |  |  |  |  |
| 15-19 | 2842 | 21.4 | 2692 | 20.4 | 3168 | 22.1 |
| 20-24 | 2591 | 19.5 | 2364 | 17.9 | 2350 | 16.4 |
| 25-29 | 2428 | 18.3 | 2260 | 17.1 | 2021 | 14.1 |
| 30-34 | 1770 | 13.4 | 2118 | 16 | 2082 | 14.5 |
| 35-39 | 1409 | 10.6 | 1561 | 11.8 | 2044 | 14.3 |
| 40-44 | 1132 | 8.5 | 1250 | 9.5 | 1481 | 10.3 |
| 45-49 | 1089 | 8.2 | 969 | 7.3 | 1203 | 8.4 |
| **Women's education** |  |  |  |  |  |  |
| No education | 2049 | 15.5 | 1641 | 12.4 | 1361 | 9.5 |
| Primary | 9086 | 68.5 | 8501 | 64.3 | 8367 | 58.3 |
| Secondary | 1934 | 14.6 | 2719 | 20.6 | 3997 | 27.9 |
| Higher | 193 | 1.5 | 353 | 2.7 | 625 | 4.4 |
| **Women's occupation** |  |  |  |  |  |  |
| No occupation | 2166 | 16.4 | 1907 | 14.4 | 3819 | 26.6 |
| Professional | 238.1 | 1.8 | 329 | 2.5 | 368 | 2.6 |
| Home occupations | 8880 | 67.1 | 9089 | 68.8 | 5296 | 36.9 |
| Other | 1957 | 14.8 | 1879 | 14.2 | 4866 | 33.9 |
| **Husband's desire for children** |  |  |  |  |  |  |
| Both want same | 3935 | 58.8 | 4112 | 60.7 | 4150 | 58.1 |
| Husband wants more | 692 | 10.3 | 812 | 12 | 1065 | 14.9 |
| Husband wants fewer | 1170 | 17.5 | 1205 | 17.8 | 1412 | 19.8 |
| Don't know | 900 | 13.4 | 644 | 9.5 | 520 | 7.3 |
| **Main source of FP information** |  |  |  |  |  |  |
| Radio | 4524 | 44 | 3291 | 37.7 | 3173 | 33.3 |
| TV | 350 | 3 | 476 | 5.5 | 714 | 7.5 |
| Newspaper/Magazine | 348 | 3 | 677 | 7.8 | 774 | 8.1 |
| FP worker | 1173 | 11 | 1365 | 15.7 | 2072 | 21.7 |
| Health Facility | 3814 | 37.4 | 2914 | 33.4 | 2804 | 29.4 |
| **Decision maker for using contraception** |  |  |  |  |  |  |
| Mainly respondent | 282 | 8 | 319 | 8.7 | 444 | 9.5 |
| Mainly husband, partner | 113 | 3 | 75 | 2.1 | 124 | 2.6 |
| Joint decision | 3092 | 88 | 3272 | 89.2 | 4103 | 87.6 |
| Other | 9 | 0.3 | 3 | 0.1 | 12 | 0.3 |
| **Knowledge of ovulatory cycle** |  |  |  |  |  |  |
| Know ovulatory cycle | 1641 | 12.4 | 2589 | 19.6 | 2370 | 16.5 |
| Don't know | 11620 | 87.6 | 10611 | 80.4 | 11980 | 83.5 |
| **Ideal number of children** |  |  |  |  |  |  |
| None or no response | 226 | 1.7 | 192 | 1.5 | 351 | 2.4 |
| 1-3 Children | 8429 | 63.6 | 80 08 | 60.6 | 7942 | 55.4 |
| Four children or above | 4607 | 34.7 | 5013 | 37.9 | 6056 | 42.2 |
| **Children ever born** |  |  |  |  |  |  |
| None | 4921 | 37.1 | 4528 | 34.3 | 5163 | 36 |
| 1-3 children | 4268 | 32.2 | 5012 | 37.9 | 5524 | 38.5 |
| Four and above | 4073 | 30.7 | 3673 | 27.8 | 3663 | 25.5 |
| **Number of living children** |  |  |  |  |  |  |
| None | 5021 | 37.9 | 4609 | 34.9 | 5223 | 36.4 |
| 1-3 children | 4833 | 36.4 | 5459 | 41.3 | 5866 | 40.9 |
| Four and above | 3408 | 25.7 | 3146 | 23.8 | 3260 | 22.7 |
| **Partner's age (in years)** |  |  |  |  |  |  |
| 29 or below | 1798 | 26.6 | 1509 | 22 | 1150 | 15.8 |
| 30-39 | 2366 | 35 | 2793 | 40.6 | 3127 | 42.8 |
| 40-49 | 1665 | 24.6 | 1669 | 24.3 | 2048 | 28 |
| 50 or above | 928 | 13.7 | 903 | 13.1 | 980 | 13.4 |
| **Partner's education** |  |  |  |  |  |  |
| No education | 1733 | 21.3 | 1555 | 18.8 | 1011 | 13.8 |
| Primary | 5383 | 66 | 5640 | 68.3 | 4900 | 67.1 |
| Secondary | 870 | 10.7 | 787 | 9.5 | 992 | 13.6 |
| Higher | 165 | 2 | 274 | 3.3 | 403 | 5.5 |
| **Partner's occupation** |  |  |  |  |  |  |
| No occupation | 101 | 1.2 | 67 | 0.8 | 567 | 7.8 |
| Professional | 282 | 3.5 | 513 | 6.2 | 412 | 5.6 |
| Home occupations | 5736 | 70.6 | 5233 | 63.4 | 2517 | 34.5 |
| Other | 2012 | 24.7 | 2435 | 29.5 | 3810 | 52.2 |
| **Number of HH members** |  |  |  |  |  |  |
| 1-5 members | 7368 | 55.6 | 7736 | 58.6 | 8520 | 59.4 |
| 6-10 members | 5666 | 42.7 | 5321 | 40.3 | 5682 | 39.6 |
| 11-22 members | 228 | 1.7 | 157 | 1.2 | 148 | 1 |
| **Religion** |  |  |  |  |  |  |
| Catholic | 5691 | 43 | 5258 | 39.8 | 5257 | 36.6 |
| Protestant | 5441 | 41.1 | 5967 | 45.2 | 6769 | 47.2 |
| Adventist | 1723 | 13 | 1571 | 11.9 | 1804 | 12.6 |
| Muslim | 171.7 | 1.3 | 256 | 1.9 | 264 | 1.8 |
| Other | 214.8 | 1.6 | 147 | 1.1 | 255 | 1.8 |
| **Sex of HH head** |  |  |  |  |  |  |
| Male | 8873 | 66.9 | 9156 | 69.3 | 9920 | 69.1 |
| Female | 4389 | 33.1 | 4058 | 30.7 | 4430 | 30.9 |
| **Marital status** |  |  |  |  |  |  |
| Single | 5112 | 38.5 | 4953 | 37.5 | 5765 | 40.2 |
| Married | 4722 | 35.6 | 4623 | 35 | 4656 | 32.4 |
| Living with partner | 2035 | 15.3 | 2284 | 17.3 | 2650 | 18.5 |
| Widowed | 713 | 5.4 | 561 | 4.2 | 381 | 2.7 |
| Divorced | 590 | 4.4 | 364 | 2.8 | 360 | 2.5 |
| No longer living together/separated | 90 | 0.7 | 429 | 3.2 | 538 | 3.7 |
| **Province** |  |  |  |  |  |  |
| Kigali city | 1542 | 11.6 | 1760 | 13.3 | 2113 | 14.7 |
| Southern Province | 3112 | 23.5 | 3139 | 23.8 | 3013 | 21 |
| Western Province | 3228 | 24.3 | 2918 | 22.1 | 3120 | 21.7 |
| Northern Province | 2220 | 16.7 | 2178 | 16.5 | 2187 | 15.2 |
| Eastern Province | 3160 | 23.8 | 3218 | 24.4 | 3917 | 27.3 |
| **Type of residence** |  |  |  |  |  |  |
| Urban | 1991 | 15 | 2548 | 19.3 | 2843 | 19.8 |
| Rural | 11271 | 85 | 10666 | 80.7 | 11507 | 80.2 |
| **Wealth index** |  |  |  |  |  |  |
| Poorest | 2535 | 19.1 | 2512 | 19 | 2701 | 18.8 |
| Poorer | 2589 | 19.5 | 2578 | 19.5 | 2719 | 19 |
| Middle | 2678 | 20.2 | 2548 | 19.3 | 2712 | 18.9 |
| Richer | 2595 | 19.6 | 2595 | 19.6 | 2899 | 20.2 |
| Richest | 2865 | 21.6 | 2980 | 22.6 | 3318 | 23.1 |
| **Own TV or Radio** |  |  |  |  |  |  |
| No TV or Radio | 4358 | 32.9 | 5348 | 40.5 | 7698 | 53.7 |
| HH has Radio | 8904 | 67.1 | 7866 | 59.5 | 6651 | 46.4 |

Most participants think that the ideal number of children in HH is between 1-3 children, 63.6 % (8429 women), 37.1% (4921women) had no child yet, and 37.9% (5021 women) had no living children. The number of HH members was mainly between 1 and 5 people, 55.6 % (7368 women) and the participants’ religion was mostly catholic, 43% (5691 women). Most of the surveyed women were catholic, belonging in the poor classes of wealth index (poorest and poorer) and owning radio in their HH, 43% (5691 women), 38.6% (5121 women), 67.1% (8904 women) respectively (Table 1).

For RDHS 2014/2015, most of the study participants were youth (15-24years), 38.3% (5056 women), with primary education, 64.3% (8501 women), with home occupations, 68.8 % (9089 women), and were not yet married/in union, 37.5% (4953 women). Majority of the study participants’ women were from eastern province, 24.4% (3218 women), living in rural area, 80.7% (10666 women). Women’s source of information on FP was mainly radio (37.7% (3291 women) and most of them do not know their ovulatory cycle, 80.4% (10611 women). The partner’s age was mostly ranging between 30 to 39 years, 40.6 % (2793 partners), and their education was mainly primary, 68.3% (5640 partners), with home occupation, 63.4% (5233 partners). The husband’s desire for children was mostly the same as one for the study participants; 60.7% (4112 husbands) and the decision for contraception use was in most of cases taken by both women and husbands, 89.2% (3272 couples) (Table 1).

Most participants considered that 1-3 children is the ideal number of children per HH, 60.6% (8008 women). Women informed that the number of their children ever born ranges mainly between one and three, 38.5% (5524 women), and their number of living children was between 1-3 children, 40.3% (5459 women). The number of HH members for the participants was mainly between 1 and 5 people 58.6% (7736 women) and 45.2% (5967 women) of them were protestant believers. Among the surveyed women, the big number was in the poor classes of wealth index (poorest and poorer), 38.5% (5090 women) and had a radio in their HHs 59.5% (7866 women) (Table 1).

For RDHS 2019/2020, study participants were mainly youth group (15-24years), had primary education, with home occupations and not yet married/in union, 38.5% (5518 women), 58.3% (8367 women), 36.9% (5296 women), 40.2% (5765 women) respectively (Table 1). The Majority of women were from eastern province, 27.3% (3917 women), living in rural area, 80.2% (11507 women). Women’s source of information on FP was mainly radio (33.3% (3291 women) and most of them do not know their ovulatory cycle, 83.5% (11980 women). The partner’s age was mostly ranging between 30 to 39 years, 42.8% (3127 partners), had education primary, 67.1% (4900 partners), with other occupations 52.2% (3810 partners). The husbands with their wives had same desire for children and the decision for contraception use were taken by both, 58.1% (4150), 87.6% (4103) respectively (Table 2).

**Table 2:** Bivariate analysis: use of modern FP use and the socio-demographic characteristics of the study participants.

| Variable | RDHS 2010 |  |  |  | RDHS 2014/2015 |  |  |  | RDHS 2019/2020 |  |  |  |
| --- | --- | --- | --- | --- | --- | --- | --- | --- | --- | --- | --- | --- |
|  | COR | 95% CI |  | p-value | COR | 95% CI |  | p-value | COR | 95% CI |  | p-value |
| **Women's Age (in years)** |  |  |  |  |  |  |  |  |  |  |  |  |
| 15-19 | 1 |  |  |  | 1 |  |  |  | 1 |  |  |  |
| 20-24 | 11 | 8.4 | 15.5 | p<0.001 | 12.3 | 9.1 | 16.6 | p<0.001 | 13 | 10.6 | 17 | p<0.001 |
| 25-29 | 32 | 23.5 | 42.8 | p<0.001 | 31 | 23 | 41.5 | p<0.001 | 31 | 24.1 | 39 | p<0.001 |
| 30-34 | 38 | 28.1 | 50.8 | p<0.001 | 38.4 | 29 | 51.6 | p<0.001 | 44 | 34.8 | 55 | p<0.001 |
| 35-39 | 37 | 27.6 | 50.8 | p<0.001 | 35.7 | 26 | 48.2 | p<0.001 | 38 | 30 | 48 | p<0.001 |
| 40-44 | 22 | 15.8 | 29.9 | p<0.001 | 27.6 | 20 | 37.7 | p<0.001 | 26 | 20.9 | 33 | p<0.001 |
| 45-49 | 8.9 | 6.3 | 12.6 | p<0.001 | 12 | 8.7 | 16.5 | p<0.001 | 11 | 8.4 | 14 | p<0.001 |
| **Women's education** |  |  |  |  |  |  |  |  |  |  |  |  |
| No education | 1 |  |  |  | 1 |  |  |  | 1 |  |  |  |
| Primary | 1 | 0.9 | 1.2 | 0.529 | 1 | 0.8 | 1.1 | 0.435 | 1 | 0.8 | 1 | 0.51 |
| Secondary | 0.6 | 0.5 | 0.7 | p<0.001 | 0.4 | 0.3 | 0.4 | p<0.001 | 0.4 | 0.4 | 1 | p<0.001 |
| Higher | 1.3 | 0.9 | 1.7 | 0.111 | 0.8 | 0.6 | 1 | 0.094 | 0.7 | 0.5 | 1 | 0.001 |
| **Women's occupation** |  |  |  |  |  |  |  |  |  |  |  |  |
| No occupation | 1 |  |  |  | 1 |  |  |  | 1 |  |  |  |
| Professional | 3.9 | 2.8 | 5.2 | p<0.001 | 5.2 | 3.9 | 6.9 | p<0.001 | 3 | 2.3 | 4 | p<0.001 |
| Home occupations | 2.8 | 2.3 | 3.2 | p<0.001 | 3.9 | 3.2 | 4.7 | p<0.001 | 2.9 | 2.6 | 3 | p<0.001 |
| Other | 1.7 | 1.4 | 2.1 | p<0.001 | 4.1 | 3.4 | 4.9 | p<0.001 | 2.8 | 2.5 | 3 | p<0.001 |
| **Knowledge of ovulatory cycle** |  |  |  |  |  |  |  |  |  |  |  |  |
| Know ovulatory cycle | 1 |  |  |  | 1 |  |  |  | 1 |  |  |  |
| Don't know ovulatory cycle | 0.1 | 0.8 | 1 | 0.178 | 1.1 | 1 | 1.3 | 0.032 | 0.9 | 0.8 | 1 | 0.1 |
| **Marital status** |  |  |  |  |  |  |  |  |  |  |  |  |
| Single | 1 |  |  |  | 1 |  |  |  | 1 |  |  |  |
| Married | 31 | 24.9 | 37.9 | p<0.001 | 24.2 | 20 | 29.2 | p<0.001 | 14 | 12.3 | 16 | p<0.001 |
| Living with partner | 24 | 19.5 | 30.2 | p<0.001 | 23.3 | 19 | 28.3 | p<0.001 | 19 | 16 | 21 | p<0.001 |
| Widowed | 2.7 | 1.9 | 3.87 | p<0.001 | 3.5 | 2.5 | 4.8 | p<0.001 | 2.1 | 1.5 | 3 | p<0.001 |
| Divorced | 7.3 | 5.3 | 9.9 | p<0.001 | 6.6 | 4.7 | 9.4 | p<0.001 | 4.7 | 3.6 | 6 | p<0.001 |
| Separated | 10 | 6.1 | 17.7 | p<0.001 | 8.7 | 6.4 | 11.8 | p<0.001 | 5.7 | 4.5 | 7 | p<0.001 |
| **Partner's age (in years)** |  |  |  |  |  |  |  |  |  |  |  |  |
| 29 or below | 1 |  |  |  | 1 |  |  |  | 1 |  |  |  |
| 30-99 | 1.4 | 1.2 | 1.6 | p<0.001 | 1.2 | 1 | 1.4 | 0.013 | 0.9 | 0.8 | 1 | 0.205 |
| 40-49 | 1.1 | 0.9 | 1.2 | 0.505 | 1.1 | 0.9 | 1.3 | 0.447 | 0.7 | 0.6 | 1 | p<0.001 |
| 50 or above | 0.6 | 0.5 | 0.7 | p<0.001 | 0.7 | 0.6 | 0.8 | p<0.001 | 0.5 | 0.4 | 1 | p<0.001 |
| **Partner's education** |  |  |  |  |  |  |  |  |  |  |  |  |
| No education | 1 |  |  |  | 1 |  |  |  | 1 |  |  |  |
| Primary | 1.6 | 1.4 | 1.8 | P<0.001 | 1.4 | 1.2 | 1.6 | p<0.001 | 1.1 | 0.9 | 1 | 0.422 |
| Secondary | 1.7 | 1.4 | 2 | P<0.001 | 1.3 | 1 | 1.6 | 0.019 | 1 | 0.8 | 1 | 0.88 |
| Higher | 1.7 | 1.3 | 2.2 | P<0.001 | 1.22 | 0 | 1.6 | 0.114 | 0.7 | 0.5 | 1 | 0.01 |
| **Partner's occupation** |  |  |  |  |  |  |  |  |  |  |  |  |
| No occupation | 1 |  |  |  | 1 |  |  |  | 1 |  |  |  |
| Professional | 1.4 | 0.9 | 2.2 | 0.186 | 0.8 | 0.4 | 1.3 | 0.34 | 0.9 | 0.7 | 1 | 0.61 |
| Home occupations | 1.2 | 0.8 | 1.8 | 0.429 | 0.9 | 0.5 | 1.6 | 0.74 | 1.4 | 1.2 | 2 | p<0.001 |
| Other | 1.4 | 0.9 | 2.1 | 0.153 | 1.1 | 0.6 | 1.8 | 0.83 | 1.3 | 1.1 | 2 | 0.01 |
| **Main source of FP information** |  |  |  |  |  |  |  |  |  |  |  |  |
| Radio | 1 |  |  |  | 1 |  |  |  | 1 |  |  |  |
| TV | 1.2 | 0.8 | 1.6 | 0.311 | 0.9 | 0.7 | 1.2 | 0.61 | 0.7 | 0.6 | 1 | 0.001 |
| News paper/Magazine | 0.9 | 0.7 | 1.2 | 0.517 | 0.7 | 0.6 | 0.9 | 0.004 | 0.4 | 0.3 | 1 | p<0.001 |
| FP worker | 2.7 | 2.4 | 3.2 | p<0.001 | 2.5 | 2.2 | 2.9 | p<0.001 | 2 | 1.8 | 2 | p<0.001 |
| Health facility | 3.9 | 3.5 | 4.4 | p<0.001 | 2.5 | 2.2 | 2.8 | p<0.001 | 2.1 | 1.9 | 2 | p<0.001 |
| **Decision maker for using contraception** |  |  |  |  |  |  |  |  |  |  |  |  |
| Mainly respondent | 1 |  |  |  | 1 |  |  |  | 1 |  |  |  |
| Mainly husband, partner | 0.8 | 0.6 | 0.9 | 0.003 | 0.4 | 0.2 | 0.9 | 0.024 | 0.4 | 0.3 | 1 | 0.007 |
| Joint decision | 1 | 0.9 | 1.2 | 0.55 | 0.6 | 0.4 | 1 | 0.05 | 0.4 | 0.3 | 1 | p<0.001 |
| Other | 0.4 | 0.3 | 0.5 | p<0.001 |  |  |  |  |  |  |  |  |
| **Husband's desire for children** |  |  |  |  |  |  |  |  |  |  |  |  |
| Wants the same | 1 |  |  |  | 1 |  |  |  | 1 |  |  |  |
| Husband wants more | 0.1 | 0.9 | 0.6 | 0.003 | 0.8 | 0.7 | 1 | 0.021 | 0.8 | 0.7 | 1 | 0.002 |
| Husband wants fewer | 0.1 | 1.2 | 0.9 | 0.55 | 1 | 0.9 | 1.2 | 0.77 | 1 | 0.9 | 1 | 0.625 |
| Don't know | 0 | 0.4 | 0.3 | P<0.001 | 0.7 | 0.6 | 0.9 | p<0.001 | 0.6 | 0.5 | 1 | p<0.001 |
| **Ideal number of children** |  |  |  |  |  |  |  |  |  |  |  |  |
| None or no response | 1 |  |  |  | 1 |  |  |  | 1 |  |  |  |
| 1-3 Children | 2.9 | 1.8 | 4.6 | p<0.001 | 1.3 | 0.9 | 1.9 | 0.159 | 1.6 | 1.2 | 2 | 0.001 |
| Four children or above | 4.1 | 2.5 | 6.6 | p<0.001 | 1.7 | 1.2 | 2.4 | 0.005 | 1.6 | 1.3 | 2 | p<0.001 |
| **Children ever born** |  |  |  |  |  |  |  |  |  |  |  |  |
| None | 1 |  |  |  | 1 |  |  |  | 1 |  |  |  |
| 1-3 children | 123 | 80.8 | 186 | p<0.001 | 209 | 120 | 363 | p<0.001 | 106 | 76.4 | 146.3 | p<0.001 |
| Four and above | 117 | 77 | 178 | p<0.001 | 198 | 113 | 346 | p<0.001 | 84 | 60.9 | 116.9 | p<0.001 |
| **Number of living children** |  |  |  |  |  |  |  |  |  |  |  |  |
| None | 1 |  |  |  | 1 |  |  |  | 1 |  |  |  |
| 1-3 children | 105 | 71.2 | 15 | p<0.001 | 173 | 105 | 286 | p<0.001 | 94 | 69.5 | 127.1 | p<0.001 |
| Four and above | 106 | 72.5 | 16 | p<0.001 | 169 | 102 | 280 | p<0.001 | 77 | 57.1 | 104.3 | p<0.001 |
| **Number of HH members** |  |  |  |  |  |  |  |  |  |  |  |  |
| 1-5 members | 1 |  |  |  | 1 |  |  |  | 1 |  |  |  |
| 6-10 members | 0.9 | 0.8 | 1 | 0.031 | 0.8 | 0.7 | 0.9 | p<0.001 | 0.7 | 7 | 1 | p<0.001 |
| 11-22 members | 0.4 | 0.3 | 0.5 | p<0.001 | 0.3 | 0.2 | 0.5 | p<0.001 | 0.4 | 0.2 | 1 | p<0.001 |
| **Religion** |  |  |  |  |  |  |  |  |  |  |  |  |
| Catholic | 1 |  |  |  | 1 |  |  |  | 1 |  |  |  |
| Protestant | 0.8 | 0.7 | 0.8 | p<0.001 | 0.9 | 0.8 | 0.9 | p<0.001 | 0.9 | 0.8 | 1 | 0.01 |
| Adventist | 1 | 0.9 | 1.2 | 0.916 | 1.2 | 1 | 1.3 | 0.014 | 1 | 0.9 | 1 | 0.78 |
| Muslim | 1.3 | 0.9 | 1.8 | 0.219 | 1.2 | 0.9 | 1.7 | 0.183 | 1.1 | 0.8 | 1 | 0.7 |
| Other | 1.2 | 0.9 | 1.7 | 0.263 | 1.3 | 0.9 | 1.8 | 0.226 | 0.9 | 0.7 | 1 | 0.64 |
| **Sex of HH head** |  |  |  |  |  |  |  |  |  |  |  |  |
| Male | 1 |  |  |  | 1 |  |  |  | 1 |  |  |  |
| Female | 0.3 | 0.3 | 0.3 | p<0.001 | 0.4 | 0.3 | 0.4 | P<0.001 | 0.5 | 0.4 | 1 | p<0.001 |
| **Province** |  |  |  |  |  |  |  |  |  |  |  |  |
| **Kigali city** | 1 |  |  |  | 1 |  |  |  | 1 |  |  |  |
| Southern Province | 1.2 | 1.1 | 1.5 | 0.011 | 1 | 0.9 | 1.2 | 0.659 | 1.2 | 1 | 1 | 0.039 |
| Western Province | 0.8 | 0.7 | 0.9 | 0.013 | 0.9 | 0.7 | 1 | 0.112 | 1.1 | 0.9 | 1 | 0.495 |
| Northern Province | 1.3 | 1.1 | 1.6 | 0.001 | 1.2 | 1 | 1.4 | 0.054 | 1.5 | 1.2 | 2 | p<0.001 |
| Eastern Province | 1.3 | 1.1 | 1.3 | 0.013 | 1.2 | 1 | 1.4 | 0.054 | 1.3 | 1.1 | 2 | p<0.001 |
| **Residence type** |  |  |  |  |  |  |  |  |  |  |  |  |
| Urban | 1 |  |  |  | 1 |  |  |  | 1 |  |  |  |
| Rural | 1.1 | 1 | 1.3 | 0.193 | 1.1 | 0 | 1.2 | 0.297 | 1.3 | 1.2 | 2 | p<0.001 |
| **Wealth index** |  |  |  |  |  |  |  |  |  |  |  |  |
| Poorest | 1 |  |  |  | 1 |  |  |  | 1 |  |  |  |
| Poorer | 0.5 | 0.9 | 1.2 | 0.613 | 1.1 | 0.9 | 1.2 | 0.521 | 0.8 | 0.8 | 1 | 0.003 |
| Middle | 2.3 | 1 | 1.4 | 0.02 | 1.1 | 0.9 | 1.2 | 0.358 | 0.9 | 0.8 | 1 | 0.041 |
| Richer | 3.9 | 1.2 | 1.5 | P<0.001 | 10 | 0.9 | 1.1 | 0.582 | 0.8 | 0.7 | 1 | p<0.001 |
| Richest | 1.2 | 0.9 | 1.3 | 0.231 | 0.8 | 0.7 | 1 | 0.017 | 0.5 | 0.4 | 1 | p<0.001 |
| **Own TV or Radio** |  |  |  |  |  |  |  |  |  |  |  |  |
| No TV or Radio | 1 |  |  |  | 1 |  |  |  | 1 |  |  |  |
| HH has TV or radio | 1.3 | 1.2 | 1.4 | P<0.001 | 1.1 | 1 | 1.2 | 0.006 | 0.9 | 0.8 | 1 | 0.004 |

### The modern FP use among women of reproductive age between 2010 and 2019

The proportion of women of reproductive age who used modern FP method increased from 24.8% in RDHS 2010, to 27.1% women for RDHS 2014/2015 and then to 33.9% women for RDHS 2019/2020. This trend was also positive and significant (Chi-square value (Chi2) = 308.5, p-value<0.001(Figure 1).

**Figure 1:**
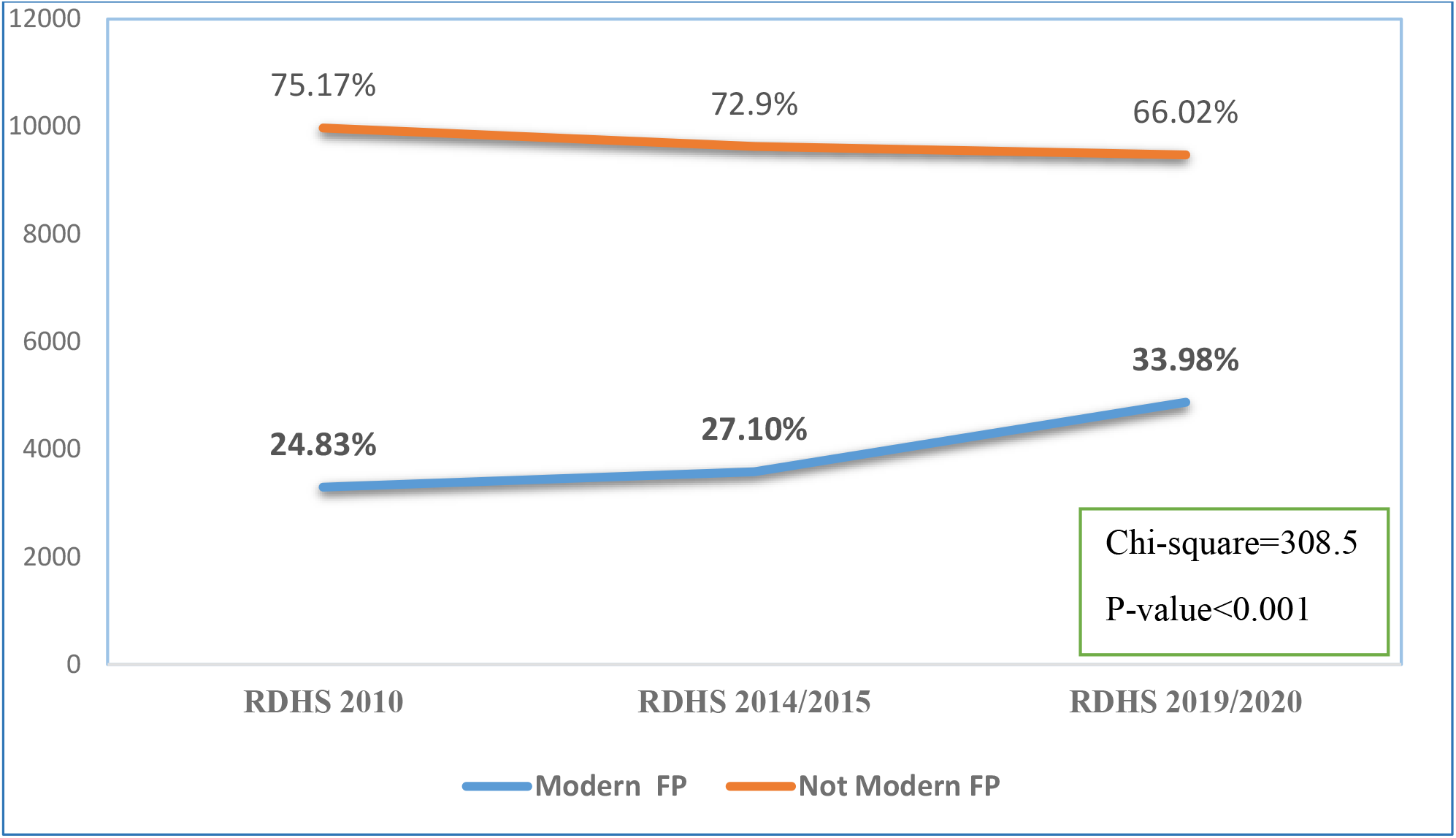
The evolution of modern FP methods use between 2010 and 2019/2020.

### Bivariate analysis: use of FP methods and socio-demographic characteristics of the study population

In all periods (2010, 2014/2015, and 2019/2020), the pairwise (unadjusted) analysis demonstrated variables that are statistically associated with the use of modern family planning in a consistent manner. These are woman’s age, women’s occupation, marital status, children ever born, and number of living children (p<0.001) (Table 2).

In addition, across the three periods, variables such as women’s education, partner’s age, partner’s education, the main source of FP information, decision maker for using contraception, partner’s age, partner’s education, husband’s desire for children, ideal number of children, number of HH members, religion, sex of HH head, residential province, and wealth index, are statistically associated with the use of modern FP methods (p<0.05). However, their significance levels varied differently within categories and periods (Table 2).

Moreover, in period of 2014/2015, only one variable (knowledge of ovulatory cycle) demonstrated a statistically significant association with use of modern family planning (Table 2).

### Factors associated with the use of modern FP methods

In 2010, a higher likelihood of using modern contraceptive methods was observed among women who reported that the ideal number of children for a couple was between 1 and 3 (aOR: 3.6; 95% CI: 1.2–10.5), those affiliated with the Muslim religion (aOR: 8.2; 95% CI: 1.0–64.4), women in a married or cohabiting relationship (aOR: 2.2; 95% CI: 1.6–3.2), and those residing in the Northern Province (aOR: 2.1; 95% CI: 1.2–3.5). Conversely, women living in households with 11 or more members, those of the Protestant faith, and those whose husbands made decisions regarding contraceptive use were less likely to use modern contraceptive methods.

In the 2014/2015 survey, women who received family planning information from a health facility were 1.6 times more likely to use modern contraception compared to those who received the information via radio (aOR: 1.6; 95% CI: 1.2–2.1). Similar to the 2010 findings, lower likelihoods of use were noted among Protestants and those living in larger households (11 or more members). Similarly, lower contraceptive use was found among women who reported being unaware of their ovulatory cycle.

In the 2019/2020 survey, modern contraceptive use was significantly higher among women aged 20 to 44 years (aORs ranging from 2.1 to 2.8) compared to those aged 15–19 and 45–49 years. Additionally, women who independently made decisions about contraceptive use were more likely to use modern methods than those whose husbands decided or made decisions jointly. Consistent with 2010 findings, women who believed that 1 to 3 children were ideal, and those in a married or cohabiting relationship, had higher odds of using modern contraception (aOR: 1.4; 95% CI: 1.0– 2.0). However, an unexpected finding was that women with higher levels of education had a lower likelihood of using modern contraceptives (Table 3).

**Table 3:**
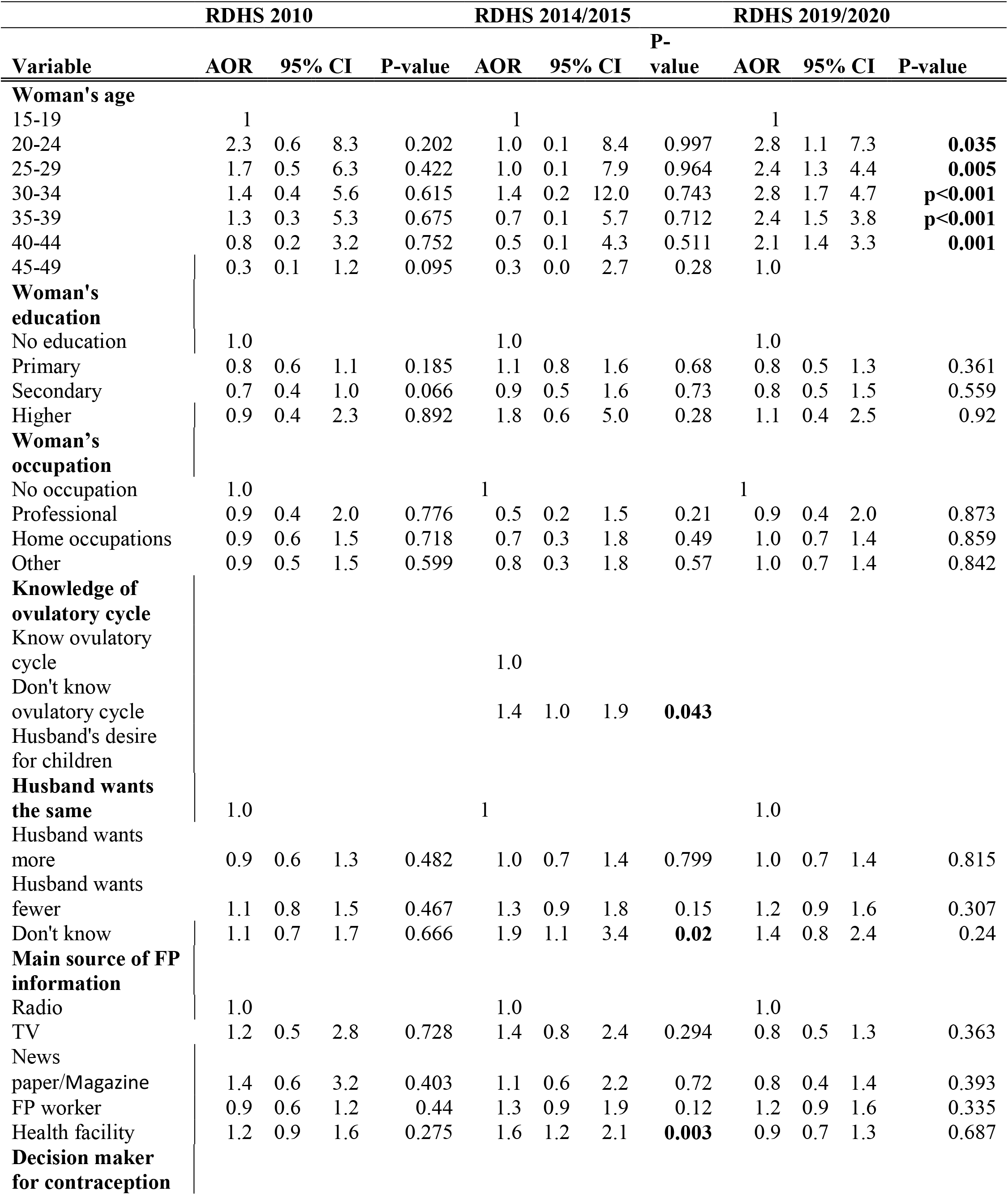
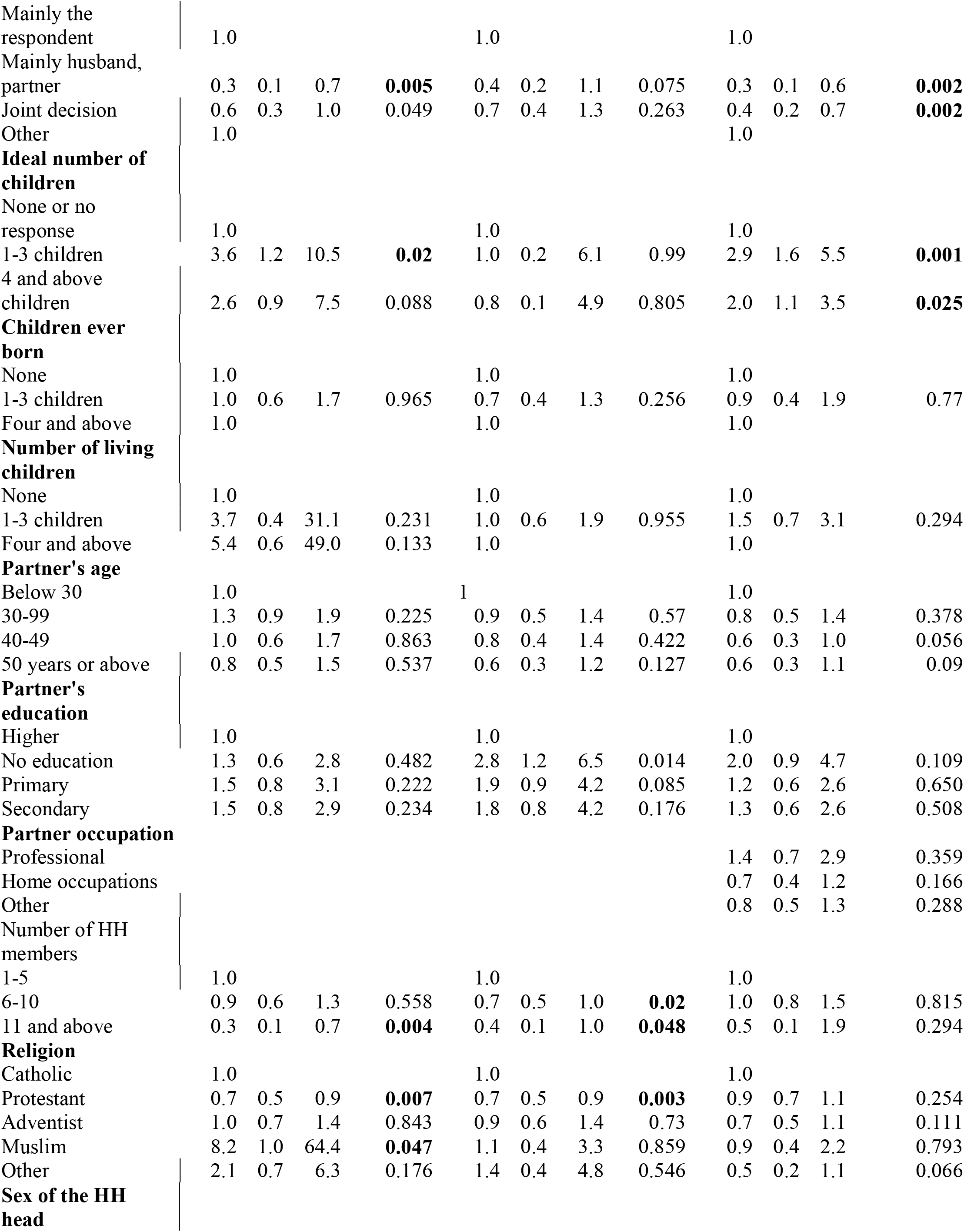
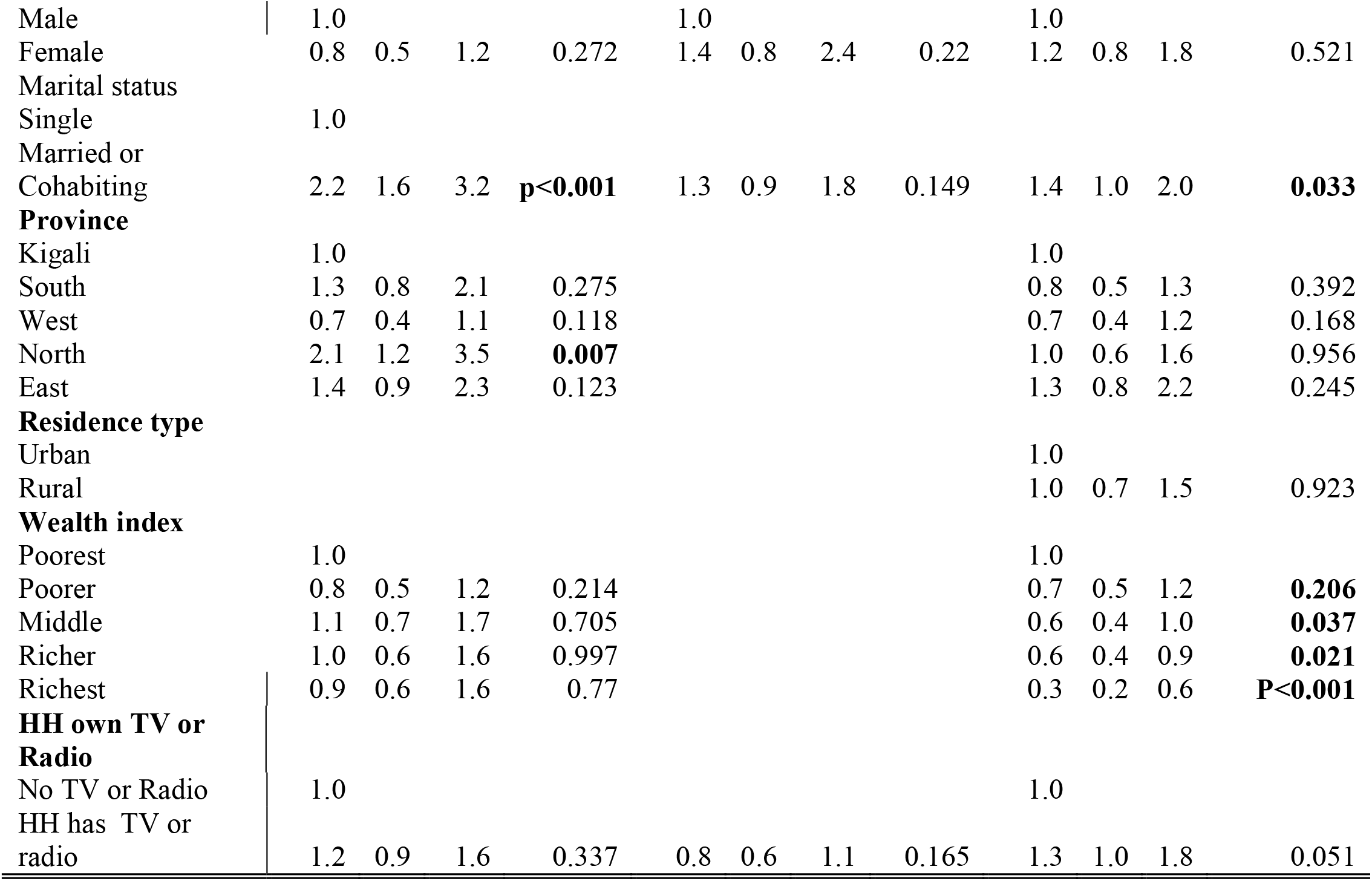
Multivariable analysis using logistic regression for Modern FP use in Rwanda.

## 4. Discussion

The gap in contraceptive behaviors among women of reproductive age can be estimated by exploring the prevalence of modern family planning and its underlying factors. The prevalence of modern FP is an effective way to track change towards reaching universal coverage in reproductive health especially the access to FP services. Trends and comparative patterns of factors linked to modern PF use across different periods needs to be established. This may contribute to the design of effective interventions focusing on the identified areas of needs. It is also important highlight strengths that may serve as examples for future designs.

The primary objective of this study was to examine the prevalence of modern family planning and its associated factors over a ten-year period using the RDHS 2010, 2014/2015, and 2019/2020. To fulfill this goal, our analysis focused on the trend of modern FP use and comparing factors underlying the prevalence of modern family planning among all women of reproductive age, across 3 different periods.

The study revealed that the use of modern FP methods increased from 25% for RDHS 2010, to 27.1% for RDHS 2014/2015 and the to 34% for RDHS 2019/2020. This steady increase may be attributed to the improved understanding of FP that is no longer a way of population control rather a tactic for empowered families (19). Similar prevalence was claimed by other studies. A study conducted in Kagera and Mara Regions of Tanzania found that the use of any modern method was estimated to 33.2 % that is slightly low when compared to our findings (20). Another study conducted in Ethiopia revealed the modern family planning method was 56.3% among 332 sexually active adult women living with HIV who visited clinics for care and treatment. This level of FP methods use was higher compared to the findings of our study(21). However, the characteristics of the populations differ at some extent, as our study considered all characteristics of women of reproductive age countrywide. A similar study conducted in Ghana claimed a progress in modern FP uptake across 2003, 2008 and 2014 DHSs. The prevalence of modern family planning was 15.7%, 18.7%, and 21,5% respectively(22). Though the study interested only in married women, this prevalence was relatively low in comparison to our findings.

Comparatively to our findings the most of reviewed studies had a low uptake of modern FP methods use. A study conducted in 17 sub-Saharan countries revealed that the contraceptive prevalence among all women aged 15-49 years was ranging from 17% to 29% (23). This study confirms that the contraceptive prevalence in Rwanda is higher than other countries in the sub-Saharan region. In the same framework, an additional study conducted in Nigerian women of reproductive age based DHS indicated that the modern FP methods use was 16.5% among those women (24), which is too low compared to our findings. Elsewhere, studies demonstrated a higher prevalence compared to our findings. In Bangladesh the prevalence of family planning use was 62.4% in 2014(25). Rodolfo et al. claimed in 2019 that in Latino America, countries like Brazil, Colombia, Costa Rica, Cuba and Paraguay have a modern contraceptive prevalence greater than 70%(26).

With regards to the factors associated with the use of modern FP methods, our analysis revealed that in the RDHS 2019/2020, women aged from 20 to 44 years were more likely to use modern FP method as compared to those aged 15-19 years. Since the FP usage coincide with the age at which Rwandan women are allowed to get married, one may conclude that adoption of modern FP methods starts when a woman get married or in union. In addition, our results for the RDHS 2019/2020 demonstrated that women living with their partners had an increased likelihood to use modern FP methods with reference to those not in union. This may be an added fact that a Rwandan woman use modern FP methods within their marriage periods. Zhihui et al. compared the contraceptive use between adolescent and adults in low- and middle-income countries from 2000 to 2017. Their findings indicated that adolescent girls were consistently less likely to use modern FP methods than adults. They were indeed more likely to have unmet needs(27). These findings concur with ours to indicate that adolescent women may not have access to modern FP methods.

The health facility as the main source of FP information (having radio as the reference category), was found to be associated with an increased likelihood of modern FP methods use according to RDHS 2014/2015. Similar results were claimed by a study carried out in Gambia using DHS data from 2013 to 2020 (28). Another study conducted in Ethiopia on fecund married women revealed that visiting a health facility was associated with an increased contraceptive use. However, this study slightly differ from ours in that it was concerned with a subpopulation, and all methods of FP were considered without distinction between modern and traditional methods(29).

The number of HH members (11-22 members versus 1-5 members) was found to be associated with less likelihood of modern FP methods’ use for RDHS 2010 and RDHS 2014/2015. There was no study found with similar results but a study conducted in Sub-Saharan Africa with a contradictory results found that the higher number of household members was associated with the use of modern FP methods (31). Since such results were not demonstrated in RDHS 2019/2020, one may say that FP community interventions have positively impacted on this group (32).

The group of women without knowledge of the ovulatory cycle were associated with improved likelihood to use modern methods of FP for RDHS 2014/2015, with reference to women knowledgeable of ovulatory cycle. Similar findings were claimed elsewhere. A study conducted in Senegal using 2014 DHS data found that knowledge of ovulatory cycle was less likely linked to modern FP use(33). The ovulation happens in the middle of the cycle so it comprehensive knowledge may also serve as an important skill to the practice of natural FP techniques that would replace the modern one(34). A study that contradicts these findings was conducted using data from 27 African countries. Its results demonstrated that poor knowledge on ovulatory cycle among adolescent women aged 15-19 years, constituted for them a strong risk factors for unwanted pregnancies(35).

Having a determined fertility preferences might predict one’s behaviors towards modern family planning (36) Our findings from 2010 RDHS and 2019/2020 revealed that women who believe that the ideal number of children is from 1 to 3 per family had the higher odds to use modern family planning as compared to women without choice. the analysis of RDHS2019/2020 revealed in addition, a positive relationship between the ideal number (4 and above vs 0 or no choice) and the modern Family planning use. This reality might be a result of previous intervention that encouraged Rwandan families to have as fewer children as possible for their development.

The residence in the Northern province with reference to the city of Kigali was revealed to be associated with less probability of using modern methods of FP for RDHS 2010. Residence region was also found to be associated with modern FP uptake in some region of India(37), and the study conducted in Nigeria that discovered a significant positive association between modern FP methods use and the region of residence among women of reproductive age of all background (24). A possible explanation is that family planning interventions may be implemented differently across the regions and provinces. Since in Rwanda most of interventions are district based, further studies should find out if disparities exist between districts.

The wealth index was also found to be associated with modern FP methods. More specifically, our study demonstrated that the middle, richer and richest wealth index compared to the poorest category were associated with the use of modern methods of FP according to analysis results of RDHS 2019/2020 data. This is an indicative that the more people become wealthier, their adherence levels to modern family planning decline. Elsewhere in Africa, same results were demonstrated: a study conducted in Ghana claimed that with reference to poorest group, women of the richest quantile were less likely to opt for modern contraceptive(22). The contradicting results were claimed by a study done in Ethiopia on married/in-union women of reproductive age using the data from DHS 2016. This study found that the middle and richer category compared to the poorest category were associated with an increased likelihood of modern FP methods uptake (29).

It is known that catholic church encourages its believers to use natural FP methods. These methods are less effective compared to modern FP methods. Our study revealed that protestant women have the low odds to use modern family planning in reference to the catholic women for both RDHS2010 and 2014/2015. Since the 2 churches host more than 80% of Rwandan people, on may conclude that religion constitutes a major hindering factor to the modern contraceptive use. A study conducted in Ghana demonstrate significant differences among churches in relation to modern contraceptive use. This study found that Christian and orthodox believers were more likely to use modern FP than Muslims(22).

## 5. Conclusion

This study revealed that there has been a consistent increase in adherence to modern FP methods among Rwandan women of reproductive age for the last decade: from 25.1% for RDHS 2010, to 27% for RDHS 2014/2015 and then to 34% for RDHS 2019/2020. This study also demonstrated factors that have a negative influence on modern family planning uptake in disparate manner. These are knowledge of the ovulatory cycle that was associated with the less use of modern methods of FP for RDHS 2014/2015; the higher level of partner’s education found be to associated with less likelihood of modern FP methods use in both RDHS 2014/2015 and RDHS 2019/2020; increase number of household members which was associated with poor likelihood to use modern FP methods, protestant religion that decreased the odds of using modern contraceptive for the 2010RDHS and 2014/2920, and the wealth index categories of middle, richer and richest which were negatively associated with the use of modern methods of FP according to RDHS 2019/2020 data. One the other hand, factors were positively associated with modern FP uptake. These include women age ranging from 20-44 years (RDHS20192020), the age category of 20-35 years at first sex (RDHS2010 and 2014/2015), women believing that ideal number of children is 1-3 (2010 RDHS) and one or more (2019/2020 RDHS), women living with their partners for (2010RDHS and 2019/2020 RDHS), residence in the Northern province that was revealed to have higher likelihood to use modern methods of FP (RDHS 2014/2015), the health facility as the main source of FP information (RDHS 2014/2015).

Based on the results of our study, it is recommended that the programs aiming at increasing the modern family planning should consider more advanced intervention targetting adolescent women, church members and rich families.

## Data Availability

The datasets can be downloaded from DHS Prograpm and STATA do file can be obtained by sending an email to the corresponding author.

https://dhsprogram.com/data/dataset_admin/index.cfm

## 6. Strengths and limitations of the study

This study used secondary data from RDHS 2010, 2014/2015 and 2019/2020 that were collected for other purposes than this study. This implies that the design, methods, and tools used for them may not fully benefits this study to avoid any kind of bias or more generally to provide all variables needed to achieve the objectives of this study. Additionally, data were collected in cross sectional manner, and the information captured verbally may not reflect the real situation of participant.

## 7. Supplementary materials

None

## 8. Acknowledgments

None

## 9. Authors’ contributions

BN, MB, ZN, JMVS, JN, MJ, IVN, HJ and CM conceptualized the study and developed a draft of the proposal. BN, ZN and MB designed the methodological approaches and analyzed the data. BN, MB, ZN, JMVS, JN, MJ, IVN, HJ and CM reviewed and improved the manuscript. All the authors contributed to the draft and final version of the paper.

## 12. Supplementary file

None

## 13. Declarations

### Ethics, approval and consent to participate in the study

This is a secondary data analysis using 2010, 2014/2015 and 2019/2020 RDHS. The participants’ electronic records were used in this study. The participants’ confidentiality, their identification and any other information that can be used to identify them, were strictly and completely secured. This study was originally a thesis for a master program in Public Health, its proposal was approved by the School of Public Health, College of Medicine and Health Sciences in the University of Rwanda. The authorization to use the dataset was issued online by the DHS Program. The data sets were stored on a secured computer using a strong password. Moreover, this study and the surveys that are the data sources were conducted *in accordance with the Declaration of Helsinki* to ensure respect, informed consent, justice, beneficence and non-maleficence in favor of participants.

### Consent for publication

Not applicable

